# Clinical Efficacy and Target Engagement of Glutamatergic Drugs: Placebo-Controlled RCTs of Pomaglumetad and TS-134 for Reversal of Ketamine-Induced Psychotic Symptoms and PharmacoBOLD in Healthy Volunteers

**DOI:** 10.1101/2020.03.09.20029827

**Authors:** Joshua T. Kantrowitz, Jack Grinband, Donald C. Goff, Adrienne C. Lahti, Stephen R. Marder, Lawrence S. Kegeles, Ragy R. Girgis, Tarek Sobeih, Melanie M. Wall, Tse-Hwei Choo, Michael F. Green, Yvonne S. Yang, Junghee Lee, Guillermo Horga, John H. Krystal, William Z. Potter, Daniel C. Javitt, Jeffrey A. Lieberman

## Abstract

We tested two metabotropic glutamate receptor 2/3 (mGluR2/3) agonist prodrugs – pomaglumetad (POMA) and TS-134 – including a high-dose of POMA that was four times the dose tested in the failed phase schizophrenia III trials – in two proof of mechanism, Phase Ib studies using identical pharmacoBOLD target-engagement methodology.

The POMA study was a double-blind, NIMH-sponsored, 10-day study of 80 or 320 mg/d POMA or placebo (1:1:1 ratio), designed to detect d>0.8 sd between-group effect-size differences. The TS-134 study was a single-blind, industry-sponsored, 6-day study of 20 or 60 mg/d TS-134 or placebo (5:5:2 ratio), designed to permit effect-size estimation for future studies. Primary outcomes were ketamine-induced changes in pharmacoBOLD in the dorsal anterior cingulate cortex (dACC) and Brief Psychiatric Rating Scale (BPRS).

95 healthy controls were randomized to POMA and 63 to TS-134. High-dose POMA had significant within and between-group reduction in ketamine-induced BPRS total symptoms (p<0.01, d=-0.41; p=0.04, d=-0.44, respectively) but neither POMA dose significantly suppressed ketamine-induced dACC pharmacoBOLD. In contrast, low-dose TS-134 had significant/trend level, moderate to large within and between group effects on BPRS positive symptoms (p=0.02, d=-0.36; p=0.008, d=-0.82, respectively) and dACC pharmacoBOLD (p=0.004, d=-0.56; p=0.079, d=-0.50, respectively) using pooled across-study placebo data.

High-dose POMA exerted significant effects on clinical symptoms, but not on target engagement, suggesting a higher dose may yet be needed. TS-134 20 mg showed evidence of symptom reduction and target engagement, indicating a curvilinear dose-response curve. These results warrant further investigation of mGluR2/3 and other glutamate-targeted treatments for schizophrenia.

## Introduction

All current FDA approved drug treatments for schizophrenia act by blocking dopamine D_2_ receptors. While they often reduce psychotic symptoms and prevent their recurrence [1,2], they have significant limitations in their efficacy and cause extensive side effects [3-6].

Efforts to develop drugs acting at novel molecular targets have been largely unsuccessful. Among the numerous candidate targets for treatment development, glutamate [7,8] represents one of the highest priorities. However, studies of glutamate-targeted drugs have not successfully led to FDA-approved treatments [9-11].

This begs the question of whether in prior studies the glutamate target itself failed, or the compounds tested failed to sufficiently engage the target. Conducting a target engagement study assesses whether the experimental agent is present in the brain and binding its molecular target in adequate concentrations to exert its therapeutic effects. While PET remains the gold-standard technology by which to measure target engagement [12], there are very few radioligands that have been developed for glutamate receptors. Other biomarkers, such as EEG or fMRI, are informative, but provide less direct, functional measures of target engagement.

Ketamine is a non-competitive antagonist at N-methyl-D-aspartate glutamate receptors (NMDAR). In animal models, ketamine administration induces an acute increase in glutamate release and recycling – sometimes termed the glutamate “surge” – that is thought to reflect an attempt of the glutamate system to achieve neurotransmission in the face of NMDAR inhibition [13-15]. Glutamate reuptake in glia accounts for ∼50% of energy expenditure in the brain [16], so that changes in glutamate/glutamine cycling and energy expenditure are highly correlated [17-20]. The increased energy expenditure, in turn, leads to an increase in local cerebral blood flow that can be detected using BOLD imaging [21-23]. As a result, the ketamine-induced changes in resting BOLD fMRI response (termed pharmacoBOLD) can be used as a putative biomarker of glutamate response [24,25] and a measure of functional target engagement.

The effects of NMDAR antagonist-induced glutamate release have been measured in rodents by microdialysis [26,27], pharmacoBOLD [21,28] and locomotor hyperactivity [29,30] and shown to be suppressed by presynaptic metabotropic glutamate type 2/3 receptor (mGluR2/3) agonists. These results should predict antipsychotic efficacy; however, they have not been replicated in humans [26,27].

We previously assessed ketamine-induced pharmacoBOLD and _1_HMRS glutamate+glutamine peak for their utility as biomarkers for subsequent target engagement studies [24]. While both were sensitive measures, pharmacoBOLD was the most robust measure, with a between group effect size of 5.4 vs. placebo. In the present project, we used pharmacoBOLD to evaluate the effects of two experimental mGluR2/3 agonist prodrugs, pomaglumetad (POMA/LY2140023) and TS-134, on ketamine effects in healthy controls. In contrast to previous single dose pharmacoBOLD studies [31], we assessed sustained exposure of these medications, more closely replicating the intended (chronic) clinical utilization.

The POMA study was funded by the National Institute of Mental Health Fast-Fail Trial for Psychotic Spectrum disorders (FAST-PS) project. We hypothesized that the reason for POMA’s failure in pivotal Phase III trials [32-34] was due to insufficient dosing and that higher doses were warranted to engage the target [31]. We compared the most efficacious POMA dose used in pivotal studies (80 mg) with a dose 4x as high (320 mg) in a multi-center study powered to detect a between group, large effect size difference (Cohen’s d≥0.80 sd). In contrast, the TS-134 study was supported by Taisho Pharmaceutical R&D Inc, and was a single-center, single-blind study using an asymmetric randomization, powered to detect within-group effects and to estimate effect-sizes to assist in the design of follow-up clinical trials.

As the studies were conducted contemporaneously using similar but not identical designs, we present the results of these two mGluR2/3 agonist studies in parallel. In particular, identical pharmacoBOLD methodology was used. Key differences included duration of dosing of experimental medication and subject monitoring (10 days of outpatient, multi-center treatment for POMA and 6 days of inpatient, single-center treatment for TS-134). As it was adequately powered, the POMA study is presented independently with primary between-group analysis, but TS-134 results are presented with primary within group, along with a post-hoc, across-study, pooled placebo group to increase the n and statistical power.

## Methods and Materials

### Subjects

Written informed consent was obtained from all participants, as approved by the participating Institutional Review Boards. Both trials were conducted under CONSORT guidelines between April 2017 and April 2019. (Clinicaltrials.gov: NCT02919774, NCT03141658).

Volunteers were aged 18 to 55 years old without current or past Axis I or II psychiatric or substance history (SCID DSM V) [35]; a first-degree relative with schizophrenia; a history of adverse reaction to an NMDAR modulator, violence, suicidality or significant medical illness, including hypertension (>140/90) or significant head injury; or current psychotropics.

### POMA study design

This was a randomized, placebo-controlled, double-blind, multisite, outpatient investigation conducted at CUMC/NYSPI, UAB, UCLA and NYULMC. Subjects were randomized in a 1:1:1 ratio to ten days of bid low-dose (80 mg/day), high-dose (320 mg/day) or placebo, with the first and last doses taken in clinic. Active POMA (LY404039) and prodrug (LY2140023) levels were assessed on day 1 (one-hour post dose), Day 5 (random) and pre/post Day 10 pharmacoBOLD scan. The planned sample size was 81 completers, powered from [31]. A randomization list was produced by the study biostatistician, using blocks of three, with stratification by site.

### TS-134 study design

This was a randomized, placebo-controlled, single-blind, single-site (CUMC/NYSPI) inpatient investigation. Study investigators and statistical/imaging analysis was operationally blinded. Subjects were randomized in a 5:5:2 ratio to six days of QD low-dose (20 mg), high-dose (60 mg) or placebo. TS-134 levels were assessed on day 6, 3 hours post dose. The planned sample size was 60 completers with valid, analyzable MRI scans, and was powered to detect within-group effects for effect-size estimation for the design of follow-up clinical trials. A randomization list was produced by the study biostatistician.

### PharmacoBOLD methods (Figure 1)

Two 15 minute T2*-weighted echoplanar imaging scans were collected before and after a 0.23 mg/kg, 1-minute bolus of racemic ketamine hydrochloride (Ketalar; Parke Davis, Morris Plains, NJ). The ketamine bolus was administered without subsequent infusion, and was identical to the bolus used in our biomarker development study [24] (see Emethods). Subjects were monitored frequently for blood pressure and continuously with EKG and pulse-oximetry by a study physician and assessed for the clearance of ketamine effects prior to discharge. A subset of subjects in both studies (n=61) underwent additional monitoring using a pulse oximeter and respiration belt.

**Figure 1:**
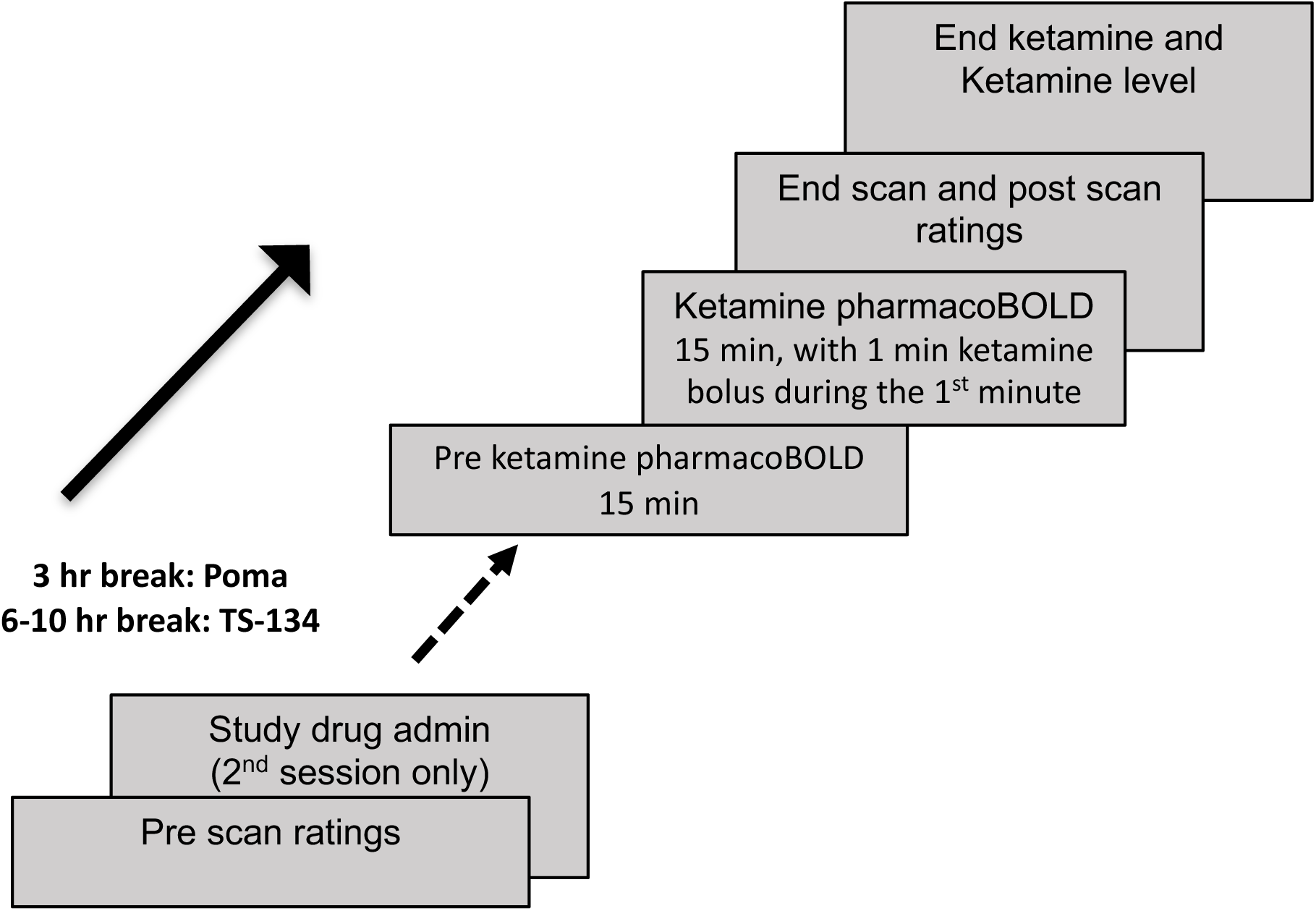
Schematic model of pharmacoBOLD sessions.

To qualify for randomization, subjects were required to show a within session increase of ≥0.5% from pre-ketamine baseline to peak ketamine response in the dorsal anterior cingulate cortex (dACC) during the screening pharmacoBOLD. The second pharmacoBOLD scan was conducted ∼3 hours after the last dose for POMA and ∼8 (6-10) hours after the last dose of TS-134, consistent with cerebrospinal fluid T_max_ of the two agents.

The Brief Psychiatric Rating Scale (BPRS) [36] was used to assess for ketamine-induced psychosis and Clinician Administered Dissociative States Scale (CADSS) [8] was used to assess for ketamine-induced non-specific disassociation. Both scales were completed twice per session: prior to and immediately following measurement of ketamine levels, approximately after MRI ended (∼20 minutes after the ketamine bolus and 5 minutes after each MRI ended.

### Regions of Interest

Based on prior work [24], our pre-defined primary ROI was the dACC. Additional voxelwise, whole brain analysis was conducted, along with secondary ROIs per a recently published study using POMA [31], including a prespecified secondary anterior insula ROI for the TS-134 study only.

### Statistical analysis

Demographics, screening pharmacoBOLD and clinical responses were summarized by randomized treatment group and differences tested using ANOVA and chi-square tests.

The study drug effect on the primary target engagement outcome (pharmacoBOLD response) was tested using linear regression models, calculated as the %difference in peak post-ketamine pharmacoBOLD response at the final session compared to screening, modeled for each treatment group. Mean within-group and pairwise contrasts for group differences were estimated and tested for significance. Models were fit both within study and in order to increase because of the asymmetric randomization and small number of participants who received placebo, using a post-hoc, pooled CUMC/NYSPI placebo group across studies for TS-134. This approach was permitted by the identical pharmacoBOLD design across studies and a comparable between study placebo group change for the BPRS total (d=-0.13) and dACC pharmacoBOLD (d=0.04). POMA models were adjusted for site. Similar models were used to assess treatment effects on clinical measurements, including the primary psychosis measure (BPRS total and subscales), the CADSS and in a post hoc subsample, controlling for heart rate in pharmacoBOLD analysis.

Spearman correlation coefficients were used to examine the association between pharmacoBOLD and symptomatic response change, and blood levels of ketamine and study drug. Analyses were carried out using SAS 9.4. Cohen’s d effect sizes, expressed in SD units, were calculated, with suppression of ketamine effects (improvements) noted by negative values.

## Results

### Ascertainment and Description of Sample (Consort charts: Supplemental Figures 1/2)

#### POMA

Ninety-five subjects were randomized in the POMA study, with 81 subjects completing both MRI scans. 76 subjects were included in the efficacy analysis, including 27 at high-dose, 21 at low-dose and 28 at placebo. Subjects were dropped from the efficacy analysis due to poor imaging quality (n=5) or noncompletion (n=14, including 6 subjects who withdrew consent, 4 adverse events, 2 lost to follow up and 2 MRI malfunction).

#### TS-134

Sixty-three subjects were randomized in the TS-134 study, with 61 subjects completing both MRI scans. 59 subjects were included in the efficacy analysis, including 25 at high-dose, 24 at low-dose and 10 at placebo. Subjects were dropped from the efficacy analysis due to poor imaging quality (n=2) or noncompletion (n=2; both subjects with adverse events unrelated to study treatment).

There were no significant within study treatment group differences at screening for either study (**Table 1**, all p>0.22).

**Table 1:** Baseline Demographics and Outcomes for Efficacy Sample^1^

|  | POMA |  |  |  |  | TS-134 |  |  |  |  |
| --- | --- | --- | --- | --- | --- | --- | --- | --- | --- | --- |
|  | Overall<br>(n=76) | Placebo<br>(n=28) | Low<br>Dose<br>(n=21) | High<br>Dose<br>(n=27) | P-values | Overall<br>(n=60) | Placebo<br>(n=10) | Low<br>Dose<br>(n=25) | High<br>Dose<br>(n=25) | P-values |
| **Age** | 34.43<br>(9.78) | 32.18<br>(8.91) | 35<br>(10.64) | 36.33<br>(9.85) | 0.28 | 38.25<br>(9.36) | 38.20<br>(7.48) | 38.28<br>(10.01) | 38.32<br>(9.52) | 1.00 |
| **% Male** | 49% | 57% | 38% | 48% | 0.42 | 68% | 50% | 76% | 68% | 0.36 |
| **dACC**<br>**Peak**<br>**Amplitude\*** | 1.90<br>(0.94) | 2.08<br>(1.29) | 1.75<br>(0.65) | 1.82<br>(0.66) | 0.41 | 2.28<br>(0.94) | 2.39<br>(1.02) | 2.34<br>(0.92) | 2.17<br>(0.96) | 0.77 |
| **BPRS**<br>**Total\*** | 3.76<br>(4.68) | 3.75<br>(4.57) | 3.71<br>(5.76) | 3.81<br>(4.00) | 1.00 | 2.32<br>(4.04) | 1.30<br>(1.95) | 2.44<br>(4.50) | 2.52<br>(4.17) | 0.70 |
| **BPRS**<br>**Positive\*** | 1.62<br>(2.05) | 1.54<br>(1.71) | 1.86<br>(2.63) | 1.52<br>(1.91) | 0.82 | 1.02<br>(1.62) | 1.00<br>(1.33) | 1.00<br>(1.55) | 1.00<br>(1.83) | 1.00 |
| **CADSS**<br>**Total\*** | 16.12<br>(14.13) | 18.86<br>(15.89) | 17.19<br>(13.14) | 12.44<br>(12.55) | 0.23 | 11.24<br>(9.61) | 9.60<br>(7.43) | 10.28<br>(7.64) | 12.48<br>(12.00) | 0.63 |

All values Mean (SD), except percentage. Abbreviations: dACC: anterior cingulate cortex; BPRS: Brief Psychiatric Rating Scale; CADSS: Clinician Administered Dissociative States Scale; \* Difference on screening day (Preinfusion vs Postinfusion). dACC amplitude represents peak change from preinfusion baseline, calculated as a percentage.
1. P-values are shown within study. There were also no significant differences for the TS-134 study when the combined placebo group was used.

### Effects on symptoms (Supplemental Table 1/2)

As expected [24], psychotic and dissociative symptoms reflected by the BPRS and CADSS ratings of subjects in all treatment groups were increased by ketamine at both screening and final sessions. The differences between the screening-final responses between treatment groups was interpreted as a measure of the POMA and TS-134 effects on ketamine stimulated glutamate neurotransmission.

#### POMA

High-dose POMA significantly reduced BPRS total scores (t_69_=2.1, p=0.04, d=-0.44, **Figure 2, top left**), along with trend-level, moderate to large effect size reductions in BPRS positive (t_69_=1.6, p=0.10, d=-0.41, **Figure 2, bottom left**) and BPRS negative (t_69_=1.9, p=0.067, d=-0.83) symptoms compared to placebo. Low-dose POMA total and BPRS positive effects were not significant, but a significant reduction vs. placebo was seen for BPRS negative (t_69_=2.2, p=0.02, d=-1.06).

**Figure 2:**
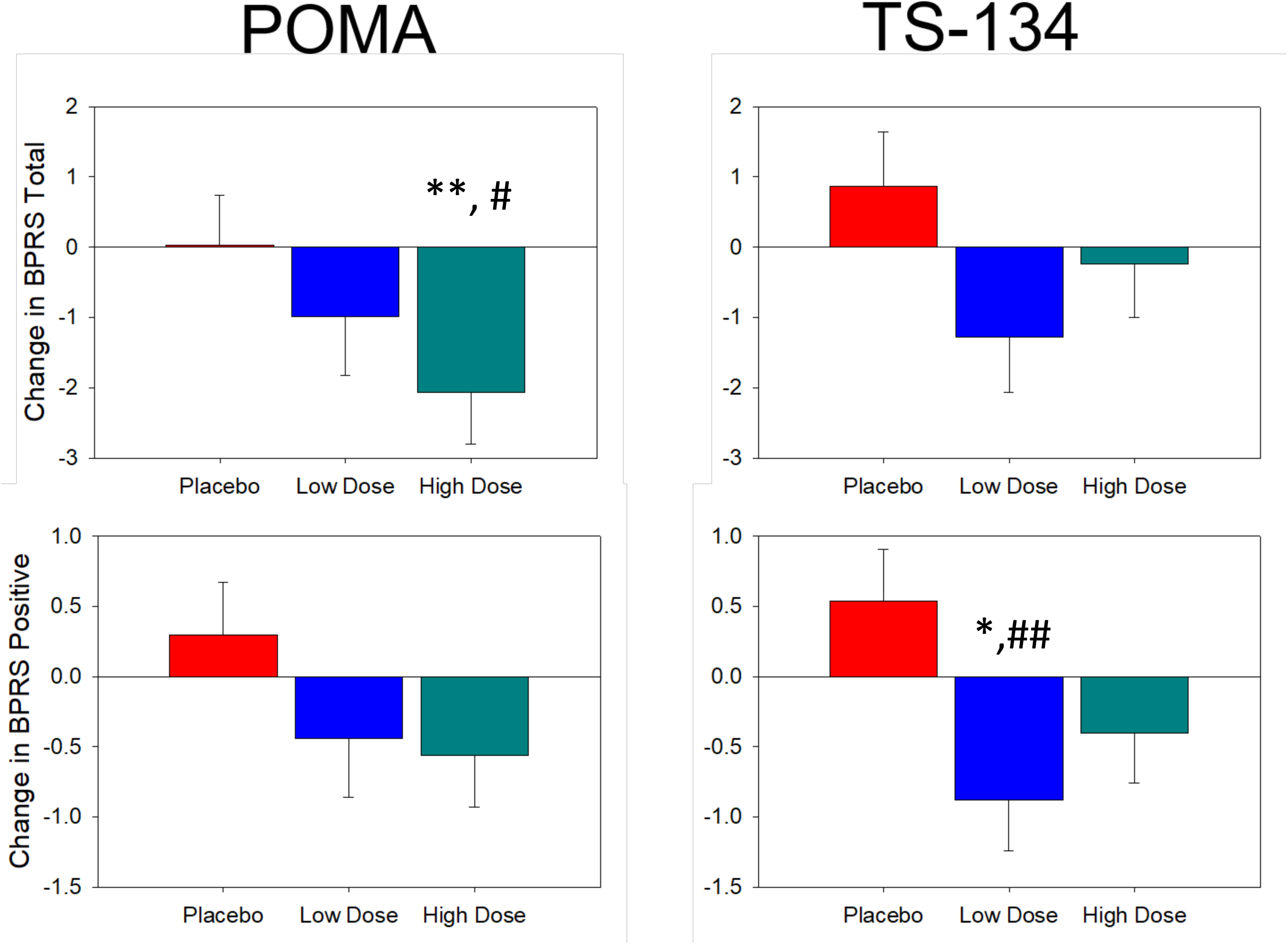
Bar graph of following ketamine administration on final assessments as compared to screening in BPRS total (top row) and BPRS positive (bottom) for the POMA and TS-134 study. TS-134 results are for the combined Columbia University Medical Center/New York State Psychiatric Institute (CUMC/NYSPI) placebo sample. Model Estimated Mean ± standard error. *=<0.05 and **=<0.01 for within group changes; #=<0.05 and ##=<0.01 for between group changes vs. placebo.

#### TS-134

Low-dose TS-134 treatment significantly reduced BPRS positive symptoms (t_56_=2.5, p=0.02, d=-0.36), along with trend-level reductions in BPRS total (t_56_=1.9, p=0.06, d=-0.32) within-groups. Because of the asymmetric randomization resulting in a small n in the placebo group of the TS-134 study, we combined subjects who received placebo from both studies to increase the n and statistical power in a post hoc analysis. The between placebo groups (POMA vs. TS-134) effect size was small for the BPRS (d=-0.13), suggesting that they were comparable and supporting the pooled placebo group analysis.

Using this pooled placebo group, we found trend-level, moderate effect size between-group differences in BPRS total symptoms (t_69_=1.9, p=0.057, d=-0.48, **Figure 2, top right**) and significant, large effect size changes in BPRS positive symptoms (t_69_=2.7, p=0.008, d=-0.82, **Figure 2, bottom right**) for low-dose TS-134 compared to placebo. High-dose TS-134 had no significant within or between-group effects.

#### CADSS

There were no significant medication effects on non-specific disassociation symptoms (CADSS) for any group in either study. Within group effects were larger in the POMA than in the TS-134 study (**Supplemental Table 1/2)**.

### Effects on MRI-pharmacoBOLD in dACC (Supplemental Table 1/2)

As previously demonstrated [24], ketamine increased BOLD activity across nearly all gray matter areas except primary visual and orbitofrontal cortex (**Fig. 3, top right**) at screening. 84.2% of 216 subjects from both studies who received a screening ketamine infusion met criteria for being a pharmacoBOLD responder to ketamine (within session increase of ≥0.5% in the dACC).

**Figure 3:**
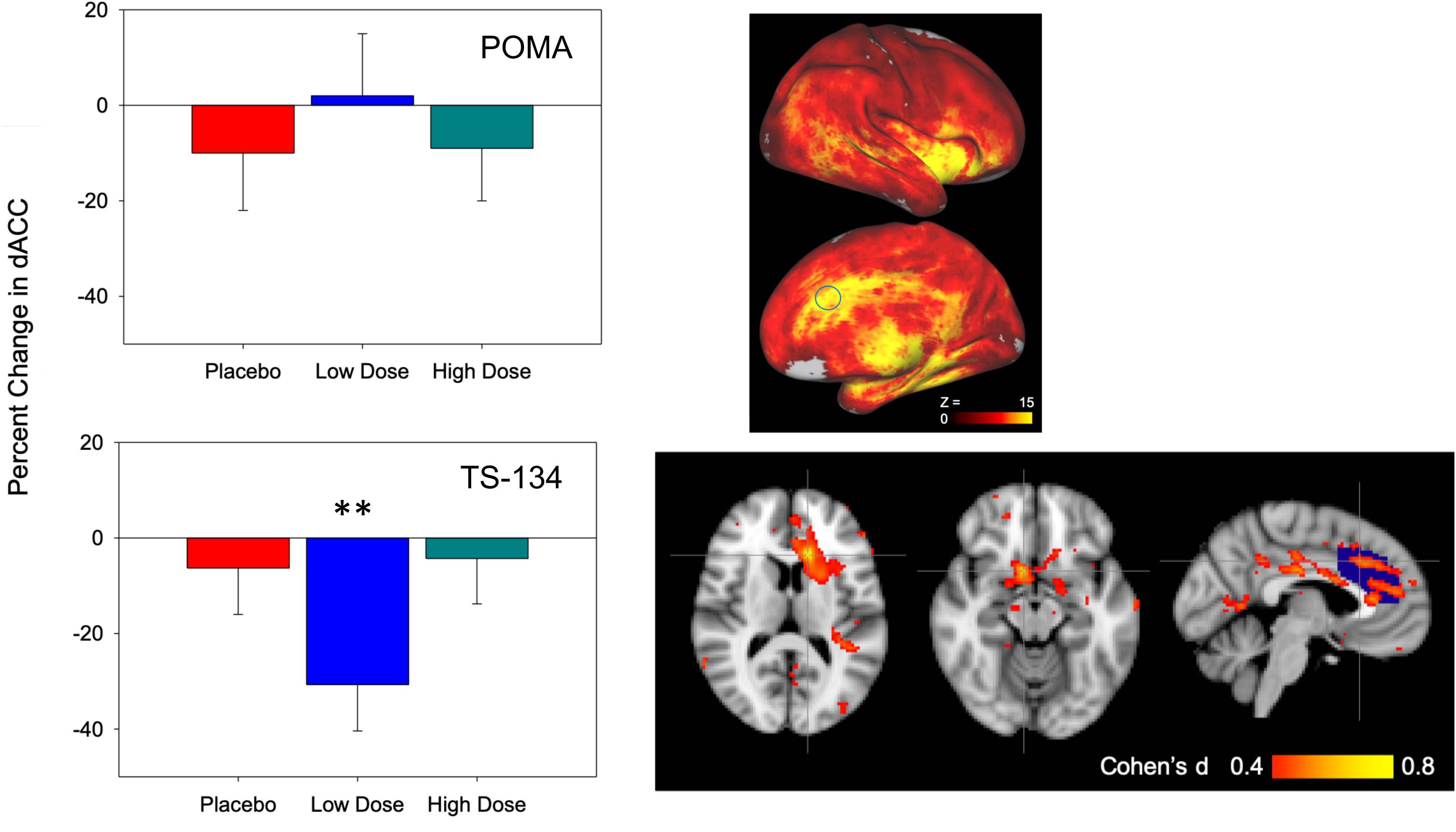
Left: Percent change in pharmacoBOLD response following ketamine administration on final assessments as compared to screening in the predefined region of interest (dorsal anterior cingulate cortex: dACC) for the POMA and TS-134 study. TS-134 results are for the combined Columbia University Medical Center/New York State Psychiatric Institute (CUMC/NYSPI) placebo sample. Model Estimated Mean ± standard error. *=<0.05 and **=<0.01 for within group changes; #=<0.05 and ##=<0.01 for between group changes vs. placebo. Right Top: Voxelwise activation maps. z Statistic maps were thresholded at z > 0. The circle indicates the region of interest used for primary analysis. Right Bottom: TS-134 20 mg dose related reduction in ketamine-evoked BOLD response. The difference in ketamine evoked BOLD suppression was compared in low dose and the combined Columbia University Medical Center/New York State Psychiatric Institute (CUMC/NYSPI) placebo sample. The thresholded effect size map (d=0.4 to 0.8) represents the contrast of (post-pre)_low_ – (post-pre)_placebo_ for 25 low dose and 23 placebo subjects. Overlay shows the dACC ROI (blue) we used for the primary outcome measure map. Significant voxels were (p<0.05) present in the right ventral striatum and left caudate.

#### POMA

Neither low-dose nor high-dose POMA had significant effects on the primary outcome measure of target engagement, dACC pharmacoBOLD response (**Fig. 3, top left**).

#### TS-134

In contrast, low-dose TS-134 produced a significant, within-group, reduction on dACC pharmacoBOLD of 30.7% (t_56_=2.98, p=0.004, d=-0.56), and a nonsignificant, moderate effect size between-group, within study difference vs. placebo of moderate-effect size (t_56_=-1.6, p=0.12, d=-0.57).

The between placebo groups (POMA vs. TS-134) effect size was small for the dACC pharmacoBOLD (d=0.04), supporting the pooled placebo group analysis. Using the pooled placebo group, trend-level differences of moderate effect size were seen (t_70_=1.8, p=0.079, d=-0.50, **Fig. 3, bottom left**) for low-dose TS-134. High-dose TS-134 had no significant within or between-group effects on dACC pharmacoBOLD.

### Correlational Analyses

There were no significant correlations between symptoms and pharmacoBOLD responses in either study, nor any significant age, site or gender effects for the main outcomes of either study.

### Additional PharmacoBOLD Analyses

We examined treatment effects on additional ROI’s that had been interrogated in a previous POMA study [31] and found a significant between-group difference in the supragenual paracingulate gyrus of moderate effect size for low-dose TS-134 (p=0.02, d=-0.66, **Supplemental Figure 3**), but neither POMA dose nor high-dose TS-134 had significant effects on any other ROIs. A significant within-group effect was observed for the prespecified secondary ROI (anterior insula) in the low-dose TS-134 group (t_55_=3.5, p=0.001, d=-0.66) but not for the high-dose group.

Finally, in the whole brain analysis, low-dose TS-134 significantly reduced BOLD increases to ketamine-induced pharmacoBOLD in the predefined dACC ROI, along with the left caudate and ventral striatum compared to the pooled placebo group (**Fig. 3, bottom right, <u>Supplemental Table 3</u>**). Whole brain analysis results were not significant after correction for multiple comparisons.

### Confounding Effects

We observed marked changes in heartrate and respiration, in response to ketamine in the course of the studies so we continuously monitored these physiologic parameters on a subset of subjects in both studies (n=61) using a pulse oximeter and respiration belt. Consistent with a prior report [37], the time course of heart rate changes mirrored that of the pharmacoBOLD response to ketamine (**<u>Supplemental Figure 4</u>**). Prompted by this, in a post hoc analysis, we examined POMA and TS-134 effects on dACC pharmacoBOLD response to ketamine controlling for heart rate and found that low-dose TS-134 exerted a moderate effect on dACC pharmacoBOLD within (d=-0.41) and between-groups (d=-0.47). Neither POMA nor TS-134 had any independent effects on heart rate or respiration.

### Pharmacokinetics (Supplemental Table 4)

Ketamine levels were consistent with previous studies [24], similar between groups for both studies, and not correlated with pharmacoBOLD/symptom responses.

POMA and TS-134 drug levels were consistent with the dose/plasma level pharmacokinetics determined in Phase 1 studies of both drugs and verified subject compliance with treatments.

### Safety

Significant nausea/vomiting was seen during early dosing for both medications, necessitating a brief dose titration schedule for both drugs (**Supplemental Figure 5/6**). Nausea/vomiting rates were greater with TS-134 than POMA. There were no serious or unexpected treatment emergent adverse events (TEAE: **Table 2)**.

**Table 2:** Safety^1^

|  | TS-134 |  |  | Pomaglumetad |  |  |
| --- | --- | --- | --- | --- | --- | --- |
| Dose (n) | 20 mg<br>(27) | 60 mg<br>(26) | Placebo<br>(10) | 80 mg<br>(24) | 320mg<br>(29) | Placebo<br>(28) |
| At least 1 TEAE | 19 (70.0%) | 16 (61.5%) | 3 (30.0%) | 15 (63%) | 19 (66%) | 10 (36%) |
| Nausea | 14 (51.9%) | 11 (42.3%) | 0 (0%) | 11 (45.8%) | 9 (31.0%) | 2 (7.1%) |
| Dizziness | 3 (11.1%) | 6 (23.1%) | 0 (0%) | 1 (4.2%) | 7 (24.1%) | 1 (3.6%) |
| Somnolence | 4 (14.8%) | 8 (30.8%) | 2 (20.0%) | 2 (8.3%) | 5 (17.2%) | 1 (3.6%) |
| Vomiting | 5 (18.5%) | 4 (15.4%) | 0 (0%) | 0 (0%) | 2 (6.9%) | 0 (0%) |
| Headache | 3 (11.1%) | 3 (11.5%) | 0 (0%) | 6 (25.0%) | 4 (13.8%) | 4 (14.3%) |
| Diarrhea | 0 (0%) | 3 (11.5%) | 0 (0%) | 0 (0%) | 3 (10.3%) | 0 (0%) |
| Pruritus/Itchiness | 0 (0%) | 2 (7.6%) | 1 (10%) | 0 (0%) | 2 (6.9%) | 0 (0%) |
| Diaphoresis | 1 (3.7%) | 1 (3.8%) | 0 (0%) | 1 (4.2%) | 2 (6.9%) | 0 (0%) |
| Dry Mouth | 0 (0%) | 1 (3.8%) | 0 (0%) | 1 (4.2%) | 0 (0%) | 3 (10.7%) |
| Weakness | 2 (7.4%) | 0 (0%) | 0 (0%) | 0 (0%) | 1 (3.4%) | 0 (0%) |

1. Treatment emergent adverse event (TEAE), n (%) reported for either study with outcome >5% within any group.

## Discussion

We believe that the results of these studies support the continued development of mGluR2/3 agonists as potential treatments for schizophrenia. Both drugs ameliorated ketamine-induced symptoms specific to schizophrenia, though only the low dose of TS-134 demonstrated target engagement reflected by attenuation of ketamine-induced pharmacoBOLD. This pattern of results can be interpreted in the following way. The high dose of POMA, though 4x the most efficacious dose used in the failed pivotal trials [32-34], may still have been too low for optimal target engagement. On the other hand, the more pharmacologically potent TS-134 achieved target engagement and symptom reduction at the low but not the high dose. Preclinical studies [29,30], indicate that TS-134 may be more potent than POMA. Moreover, the side effect rates and severity of nausea and vomiting, were also consistent with this interpretation. This pattern of results suggests that high dose POMA was on the lower part of the ascending limb of the dose response curve and the TS-134 low dose was at the asymptote while the high dose was on the descending limb of a curvilinear dose-response curve.

We examined the question of a curvilinear dose response curve for mGluR2/3 agonists by reviewing the literature and consulting with Dr. Darryl Schoepp (who developed the series of mGluR compounds including POMA as VP for Neuroscience Research at Lilly 1987-2007). There are several factors that could be result in a curvilinear dose response curve. Receptor desensitization could occur at higher occupancy levels [38], particularly at mGluR3s. High-dose POMA may not have reached the level to cause desensitization or there may have been differences between the two drugs in their proportional receptor affinities at the doses used. As previously reviewed [39], mGluR3 activation may be neuroprotective via astrocyte induced neurotrophic factors, including transforming-growth factor-β1 or glial cell-derived neurotrophic factor [40-42]. A similar curvilinear dose response for mGluR2/3 agonists has been seen in primate studies [43].

The use of pharmacoBOLD as the biomarker for target engagement also may have contributed to insensitivity and uncertainty in detecting drug effects. pharmacoBOLD provides an indirect functional measure of target engagement as opposed to PET ligands which bind directly to the molecular target. _1_H MRS may have provided a less robust but more specific measure of target engagement reflected by glutamate-glutamine concentrations in an ROI. In addition, pharmacoBOLD is also potentially confounded by cardio-respiratory effects that influence cerebrovascular blood flow [37], which may limit test-retest reliability [25].

Consequently, it will behoove investigators for future studies to consider the possible sources of the BOLD signal. Toward this end, a methodologic refinement that will improve future studies is the measurement of cardiovascular activity and respiration and controlling for their effects. In doing so, the remaining BOLD activity can more reliably be assumed to derive from brain and reflect drug effects on glutamatergic, neural activity [44-46].

Nevertheless, despite the nonspecific perfusion effects, we believe our results reflect mGluR2/3 agonist target engagement, as we demonstrated moderate effect sizes of TS-134 after controlling for heartrate (d>0.4). Our interpretation is consistent with a recent report [37] showing that ketamine produces a measurable pharmacoBOLD response after correcting for the physiologic response, and preclinical studies showing that the tachycardic effects of NMDAR antagonism are brain mediated [47-53], potentially via glutamate [48,50-53]. Moreover, NMDAR antagonism leads to similar pharmacoBOLD-like or CBV changes in both anesthetized, ventilated [54-57] and free-moving animals [26,58-61] and similar ketamine-induced effects are observed in humans across multiple imaging modalities including approaches such as _13_C MRS and PET FDG that are not affected by overall brain perfusion [37,62-64]. This argues that cardiorespiratory effects are not driving the pharmacoBOLD signal, but may add “noise” to phBOLD comparisons, and confirms that the model can still be a sensitive measure of target engagement even without adjusting for nonspecific perfusion effects. These results notwithstanding, further optimization of the approach seems warranted, including parametric variation in ketamine dose, increased physiological monitoring and statistical modeling of whole brain BOLD effects [37].

These results also confirm the reason why the 80 mg/d dose of POMA failed to separate from placebo in pivotal studies[32-34], as that dose neither blocked the ketamine-induced increases in psychotic symptoms nor pharmacoBOLD activity, indicating inadequate target engagement.

Because the TS-134 study was primarily designed to detect within-group effects to define optimal doses, an asymmetric randomization schedule was employed. In general, Cohen’s d≥0.5 are considered “visible to the naked eye of the careful observer” [65] and are widely considered to be the threshold for meaningful clinical effect. Nonetheless, while TS-134 20 mg produced significant within-group improvements on ketamine-induced positive symptoms (p=0.02) and pharmacoBOLD (p=0.004), the within study between group effects for pharmacoBOLD, though of moderate effect size (d=-0.57, **Supplemental Table 2**), were not significant. For this reason, we performed the analysis with the pooled-placebo group, which revealed a more robust between-group effect on symptoms (p=0.008), while the pharmacoBOLD effect remained at a trend-level significance, but a moderate effect size (d=-0.50). The whole brain analysis showed drug effects primarily within the predefined ROI, along with significant clusters within dopaminergic areas (ventral striatum and caudate), consistent with preclinical studies suggesting broad interactions between mGluR2/3 and dopaminergic neurotransmission [66-68]. We caution that the whole brain analysis was not corrected for multiple comparisons and requires prospective replication.

Interestingly, neither POMA nor TS-134 had effects on the CADSS ratings. This is revealing in that while the BPRS measures psychopathology, the CADSS largely measures more non-specific ketamine effects, such as disassociation, novelty and sensory perception alterations. In this context, while ketamine stimulates both psychopathology and disassociation effects, mGluR2/3 agonists act on the former and not the latter, suggesting that its effects were specifically antipsychotic.

The originally proposed trial designs called for patients to start at the target dose of their treatment group. However, because of nausea/vomiting that occurred rapidly after first dose administration, a titration schedule was instituted so that target dose in the high groups was not achieved for several days (**Supplemental Table 5/6**). The gradual titration reduced the frequency and severity of nausea substantially and no subjects discontinued due to nausea. Overall, 89.8% of the randomized sample completed the study.

Interestingly, nausea/vomiting was more severe with TS-134 (**Table 2**), supporting the increased potency of TS-134 relative to POMA. The rapid onset of nausea/vomiting suggests that the critical receptors may be either peripheral (in the gut) or in parts of the brain (e.g. brainstem/area postrema) that are not protected by the blood-brain barrier. If so, it suggests that pairing a centrally penetrant mGluR2/3 agonist with a non-penetrant antagonist may permit more rapid treatment implementation. Alternatively, a slow titration, as used here, may be required during clinical utilization.

In conclusion, our results demonstrate proof of principle and mechanism in support of the continued development of mGluR2/3 agonists as treatments for schizophrenia at optimized doses. Our negative results for POMA 80 mg/d replicate prior negative pharmacoBOLD studies [31] and offer a possible explanation for why the pivotal studies [32-34] conducted by Lilly were negative. Given modest pharmacoBOLD effects even of the highest tested dose, future development of POMA may require even higher doses. At the same time, the curvilinear dose response curve and results of the TS-134 study suggest that it is ready for a proof of concept phase IIA study at 20 mg. Future studies may be needed to refine the dosing range for other mGluR2/3 agonists as well as to better elucidate their pharmacodynamic mechanism of action.

## Data Availability

Data for FAST study is available at https://nda.nih.gov

## Acknowledgements

The POMA study was supported by the FAST-PS contract (HHSN271201200007I) from NIMH. The TS-134 study was supported by Taisho Pharmaceutical R&D Inc.

Dr. Kantrowitz reports having received consulting payments within the last 24 months from Krog & Partners Incorporated, IQVIA, Alphasights, Charles River Associates, Putnam Associates, Third Bridge, Piper Jaffray, MEDACorp, Simon Kucher, LifeSci Capital, ECRI Institute, System Analytic and BVF Partners. He has served on the Aristada Schizophrenia Advisory Board for Alkermes and the MedinCell Psychiatry Advisory Board. He has conducted clinical research supported by the NIMH, Sunovion, the Stanley Foundation, Takeda, Taisho, Lundbeck, Boehringer Ingelheim, NeuroRX, Teva and Lilly within the last 24 months. Dr. Kantrowitz a co-investigator on a study that receives lumeteperone and reimbursement for safety testing for an investigator-initiated research from Intra-Cellular Therapies Inc. He owns a small number of shares of common stock from GSK.

Dr. Goff does not accept any personal financial remuneration for consulting, speaking or research activities from any pharmaceutical, biotechnology, or medical device companies. He has received funding for research from Avanir Pharmaceuticals which does not contribute to his compensation. He has served as a consultant or advisory board member for the Welcome Trust, Intra-Cellular Therapies, Takeda, and Avanir Pharmaceuticals for which he receives no personal remuneration.

Dr. Marder has served on advisory boards for Sunovion, Roche, Boeringer Ingellheim, and Merck. He has received research support from Takeda and Boeringer Ingelheim.

Dr. Girgis reports research support from BioAdvantex, Genentech, Otsuka and Allergan and receives royalties from Routledge/Taylor and Francis.

Dr. Green has been a consultant for Biogen, Click Therapeutics, Lundbeck, and Roche, and he is on the Scientific Board of Cadent.

Dr. Krystal reports having received consulting payments from AstraZeneca Pharmaceuticals, Biogen, Biomedisyn Corporation, Bionomics, Boehringer Ingelheim International, COMPASS Pathways, Concert Pharmaceuticals, Epiodyne, EpiVario, Heptares Therapeutics, Janssen Research & Development, Otsuka America Pharmaceutical, Perception Neuroscience Holdings, Spring Care, Sunovion Pharmaceuticals, Takeda Industries, and Taisho Pharmaceutical. He has served on advisory boards for Bioasis Technologies, Biohaven Pharmaceuticals, BioXcel Therapeutics, BlackThorn Therapeutics, Cadent Therapeutics, Cerevel Therapeutics, EpiVario, Eisai, Lohocla Research Corporation, Novartis Pharmaceuticals Corporation and PsychoGenics. He is a co-sponsor of a patent for the intranasal administration of ketamine for the treatment of depression and for the treatment of suicide risk that was licensed by Janssen Pharmaceuticals; has a patent related to the use of riluzole to treat anxiety disorders that was licensed by Biohaven Pharmaceuticals; has stock or stock options in Biohaven Pharmaceuticals, Blackthorn Therapeutics, Luc Therapeutics, Cadent Pharmaceuticals, Terran Biosciences, Spring Healthcare, and Sage Pharmaceuticals. He serves on the Board of Directors of Inheris Pharmaceuticals. He receives compensation for serving as editor of the journal Biological Psychiatry.

Dr. Javitt reports having received consulting payments from Pfizer, FORUM, Autifony, Glytech, Lundbeck, Concert, and Cadence. He holds intellectual property rights for use of NMDA modulators in treatment of neuropsychiatric disorders. He holds equity in Glytech, AASI, and NeuroRx, and serves on the advisory board of Promentis, Phytec and NeuroRx.

Dr. Lieberman does not accept any personal financial remuneration for consulting, speaking or research activities from any pharmaceutical, biotechnology, or medical device companies. He receives funding and medication supplies for investigator-initiated research from Denovo, Taisho, Pfizer, Sunovion, and Genentech, and for company sponsored phase II, III, and IV studies from Alkermes, Allergan, Boehringer Ingelheim and Sunovion which does not contribute to his compensation. He receives medication supplies for investigator-initiated research from Intra-Cellular Therapies Inc. He is a consultant or advisory board member of Intra-Cellular Therapies, Takeda, Pierre Fabre, Karuna, Pear Therapeutics, Systems-1, and Psychogenics for which he receives no remuneration. He is a paid consultant for Bracket, a clinical research services organization, and holds a patent from Repligen that yields no royalties.

Dr. Grinband, Dr. Kegeles, Dr. Yang, Dr. Lee, Dr. Sobeih, Dr. Lahti, Dr. Horga, Dr. Potter, Dr. Wall and Mr. Choo reports no financial relationships with commercial interests.

